# Infant RSV immunoprophylaxis changes nasal epithelial DNA methylation at six years of age

**DOI:** 10.1101/2021.04.06.21254883

**Authors:** Cheng-Jian Xu, Nienke M. Scheltema, Cancan Qi, Rolf Vedder, Laura B.C. Klein, Elisabeth E. Nibbelke, Cornelis K van der Ent, Louis J. Bont, Gerard H. Koppelman

## Abstract

**Background:** Respiratory syncytial virus (RSV) infection has been associated with childhood wheeze and asthma, and potential mechanisms include persistent epigenetic effects.

**Methods:** In the randomized, placebo-controlled MAKI trial, 429 preterm infants randomly received RSV immunoprophylaxis with palivizumab or placebo during their first RSV season. Children were followed until age 6 for asthma evaluation. DNA methylation in cells obtained by nasal brushes at age 6 was measured by Illumina MethylationEPIC array.

**Results:** RSV immunoprophylaxis in infancy had significant impact on global methylation patterns in nasal cells at age 6. The first principal component related to the immunoprophylaxis intervention was enriched for the pathway “positive regulation of defense response to virus by host” and “antigen processing and presentation” and driven by methylation changes in *NOD2, DGKG, MSH3*, and *ITPR2*. Three CpGsites, cg18040241, cg08243963, cg19555973 were differentially methylated at genome-wide significance, but were not associated with asthma. Differential methylation region analysis identified regions near genes that were previously implicated in the development of asthma and allergy such as *HLA-DPA1, HLA-DPB1, FASLG*, and *CHI3L1*.

**Conclusions:** The study provides the first proof of concept that RSV immunoprophylaxis during infancy has long-term effects on nasal epigenetic signatures at age 6, relating to host antiviral defense pathways.

## Background

Human Respiratory syncytial virus (RSV)-related acute lower respiratory tract infection is a leading cause of severe respiratory morbidity and mortality in children [1]. RSV infection in early life is associated with increased risk of early childhood wheezing [2][3][4][5][6]. Recurrent wheezing and asthma are thought to result from alterations in early life immune development following RSV infection, when the immune system is immature. Premature born children are at higher risk for the adverse effects of early life RSV lower respiratory tract infection. However, how RSV infection would have a long-term effect on the host immune system and how this would relate to the development of asthma remains unknown.

Epigenetics refers to (heritable) changes in gene expression that are not encoded by genetic variation. Recent epigenome-wide association studies (EWAS) of allergic diseases have reported encouraging results [7][8][9][10], and epigenetics might explain the high degree of plasticity of the immune response throughout life [11]. For example, in a recent EWAS, we reported 14 CpG methylation sites in whole blood were associated with childhood asthma, that related to distinct gene expression signatures reflecting early life activation of eosinophils and effector/memory CD8 T cells and NK cells at the expense of naïve T cell subsets, providing support for a role of early life antiviral immunity in asthma inception [7]. Indeed, RSV infection may have epigenetic consequences. RSV infection of Dendritic Cells leads to alterations in histone methylation by the H3K4 demethylase KDM5B, resulting in decreased pro-inflammatory cytokine production and a subsequent increase of Th2 cytokines from T cells[12]. Moreover, RSV has been related to a nasal airway microRNA profile which is predicted to have a downregulated NF-kB signaling pathway [13]. Among these epigenetic markers, DNA methylation provides a stable marker which is suitable for investigating mechanisms for long-term regulation of gene expression upon environmental stimulation [14]. Many studies relate early life exposures to DNA methylation signatures later in life, yet these observational studies may subject to confounding and causation remains to be verified by future studies. A possible solution to this would be to nest epigenetic analyses in a randomized controlled clinical trial, where the exposure can be controlled by the randomized intervention.

We hypothesized that RSV infection in early life causes changes in DNA methylation in the nasal airway epithelium in childhood. To test this hypothesis, we investigated the randomized clinical trial MAKI [2] (ISRCTN registry, number ISRCTN73641710) to investigate the direct link between RSV prophylaxis during infancy and DNA methylation changes in the nasal epithelium at age 6. In this trial, we observed significant protective effects of early life RSV prophylaxis on having RSV infection and asthma symptoms in the first years of life [2][3]. We performed an epigenome-wide association study (EWAS) to identify differentially methylated patterns in relation to palivizumab prevention at the global, individual and regional CpG site level.

## Method

### Study design and participants

In the MAKI trial, 429 otherwise healthy late preterm infants were enrolled between 2008 and 2010, and they were born at 33 to 35 weeks of gestation. Infants were randomly assigned to receive palivizumab (n=214) or placebo (n=215) during their first RSV season. Baseline characteristics at randomization are available at NEJM.org[2]. Details about the design, definitions, protocol of the primary study, follow-up study and clinical assessment have been previously described[2][3]. Clinical assessment at age 6 years included questionnaires for respiratory symptoms (parent-reported current asthma) as well as a parent report of a physician’ s diagnosis of asthma, lung function tests (FEV_0.5,_ FVC), and assessment of specific IgE to a panel of aeroallergens as well as total serum IgE. Positive specific IgE was defined as an allergen-specific IgE concentration of at least 0·35 kU/L for house dust mite, birch, mugwort, timothy, Aspergillus, fumigatus, dog, and cat[3].

### Nasal DNA methylation measurements

Nasal epithelial cells were collected by nasal brushing around age 6 years of age at a home visit by a trained investigator. Briefly, the right nostril of the subjects was examined and the inferior turbinate was located using a speculum and penlight. The lateral area underneath the inferior turbinate was then brushed for 3 seconds with two brushes (Copan, 56380CS01 FLOQswabs) and these were placed in a 2 ml screw-cap Eppendorf tube and put into a freezer at – 80°C until further processing.

DNA was extracted from nasal brushes using the DNA investigator kit (Qiagen, Benelux BV, Venlo, the Netherlands). This was followed by precipitation-based purification and concentration using GlycoBlue (Ambion). 500 ng of DNA was bisulphite-converted using the EZ 96-DNA methylation kit (Zymo Research), following the manufacturer’s standard protocol. After verification of the bisulphite conversion step using Sanger Sequencing, DNA concentration was normalized and the samples were randomized to avoid batch effects. One standard DNA sample per chip was included in this step for quality control. In total, 296 nasal epithelium samples with sufficient DNA quality and quantity were hybridized to the Infinium HumanMethylationEPIC BeadChip array (Illumina, San Diego, CA, USA).

### Data pre-processing and statistical analysis

DNA methylation data were pre-processed in R with the Bioconductor package Minfi[15], using the original IDAT files extracted from the HiScanSQ scanner. We implemented sample filtering to remove 6 bad quality samples (call rate <99%) and 16 samples with gender mismatch. During processing, bad quality probes which failed more than 10% of the samples, the probes on sex chromosomes, the probes that mapped to multiple loci, and the probes containing SNPs at the target CpG sites with a MAF>5% in European populations were excluded^14^. We subsequently implemented stratified quantile normalization[17]. To remove the effect of extreme outliers in data, we trimmed the methylation set using: (25th percentile −3*IQR) and (75th percentile+3*IQR), where IQR=interquartile range. In our predefined analysis plan, we followed the trial design by relating the DNA methylation signatures at age 6 years to the palivizumab intervention. The association of DNA methylation with palivizumab intervention was conducted using robust linear regression corrected for sex, age at nasal brush sampling, maternal smoking, batch and surrogate variable analysis (SVA)[18] which represent unknown confounders such as cell type heterogeneity. Principal component analysis (PCA) and nonmetric multidimensional scaling (NMDS)[19] were utilized to check the global effects of palivizumab prophylaxis. R package vegan was used for nonmetric multidimensional scaling analysis (NMDS). Principal components associated with the intervention were analyzed, and the top 20 CpG sites contributing to the first two principal components were annotated using illumina Infinium MethylationEPIC Manifest File (version 1.0), with significant pathways detected by gene network (www.genenetwork.nl)[20]. For single CpG site analysis, adjustment for multiple testing was done using Benjamini & Hochberg method. Significant differentially methylated CpG site were considered when the P-value after False Discovery rate correction (FDR) was less than 0.05. For regional analysis, comb-p[21] was used to identify differentially methylated regions (DMRs) by using the summery statistics from models of single CpG site analysis. All statistical analyses were performed using the computing environment R (version 3.32).

## Results

### Sample characteristics

Figure 1 illustrated the flow chart of initial enrollment, follow-up and quality control of DNA methylation data in the MAKI study. After quality control, 274 (63.8%) of the collected samples and 790,437 probes remained for further analyses. The baseline characteristics of 274 participating children (132 palivizumab; 142 placebo), representing 63.9 % of the original study population, are presented in Table 1. Median age at the time of nasal brush collection was 6.6 years. Current asthma was defined as parent reported wheeze or asthma medication in the past 12 months[3] ; current allergic asthma was defined as having asthma and specific IgE to aeroallergens at age 6 years. Asthma prevalence was somewhat lower in the palivizumab group compared to the placebo group (14 vs 24%, *P*=0.065) (Table 1), and no differences were observed in lung function (FEV_0.5_) and total serum IgE levels, as reported previously in the full study population[3].

**Table 1.** Sample characteristics from MAKI methylation study.

|  | Palivizumab | Placebo | P-value |
| --- | --- | --- | --- |
| Sample size | 132 | 142 |  |
| Age of phenotyping (years)* | 5.79 (0.20) | 5.84 (0.28) | 0.098 |
| Age of nasal brush sampling (years) | 6.61 (0.39) | 6.65 (0.41) | 0.397 |
| Male sex, n (%) | 84 (63.6) | 69 (48.6) | 0.017 |
| Asthma , n (%) | 19 (14.4) | 34 (23.9) | 0.065 |
| FEV<sub>05</sub> (mean(sd)) | 89.51 (9.61) | 90.06 (11.49) | 0.68 |
| Total IgE (mean (sd)) | 148.70 (290.87) | 134.23 (177.47) | 0.692 |
| Total RSV infection (%) | 7 (5.3) | 22 (15.8) | 0.01 |
| Medically attended RSV infection (%) | 2 (1.5) | 13 (9.4) | 0.011 |
| RSV infection without medical attention (%) | 5 (3.8) | 9 (6.6) | 0.461 |

Abbreviation: Fev<sub>05</sub> (Forced Expiratory volume in 0.5s). IgE (Immunoglobuline E).
Missing values were not included. P-values for categorical variables were calculated using $\chi^2$ test comparing the palivizumab group and placebo group, and continuous variables by t-test.
\* the age of phenotyping of asthma, FEV<sub>05</sub> and Total IgE.

**Figure 1.**
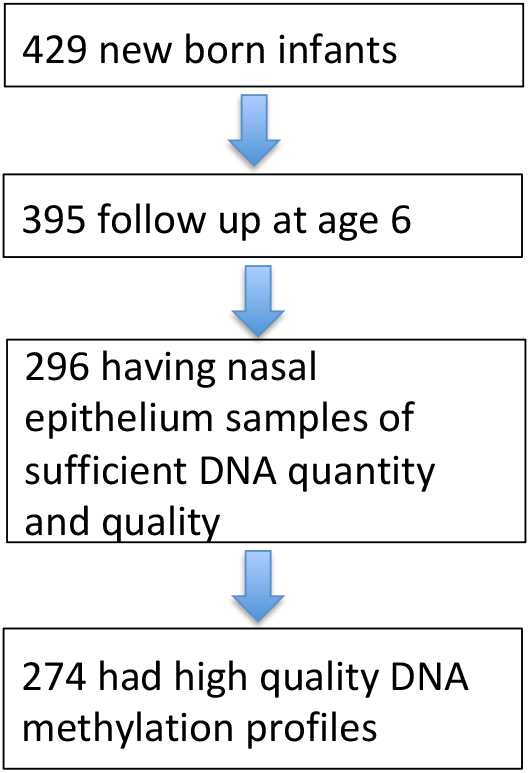
Flow chart of enrollment at age 0, follow up at age 6, and quality control of DNA methylation data in MAKI study.

### Differential methylation profiling analysis

After QC, 790,437 CpG sites were available for downstream analyses. The principal components analysis (PCA) plot (Figure 2a) and nonmetric multidimensional scaling (NMDS) plot (Figure 3) illustrated an overall methylation difference at age 6 years between the RSV immunoprophylaxis group and placebo group. The first two principal components (Pvalue_pc1_=8.1×10^−3^; Pvalue_pc2_=1.0×10^−2^) were significantly associated with RSV immunoprophylaxis (Table E1). NMDS confirmed this overall methylation difference between the two groups (adonis, R^2^=0.107, *P*=0.0055, with 1999 permutations) (Figure 3).

**Figure 2.**
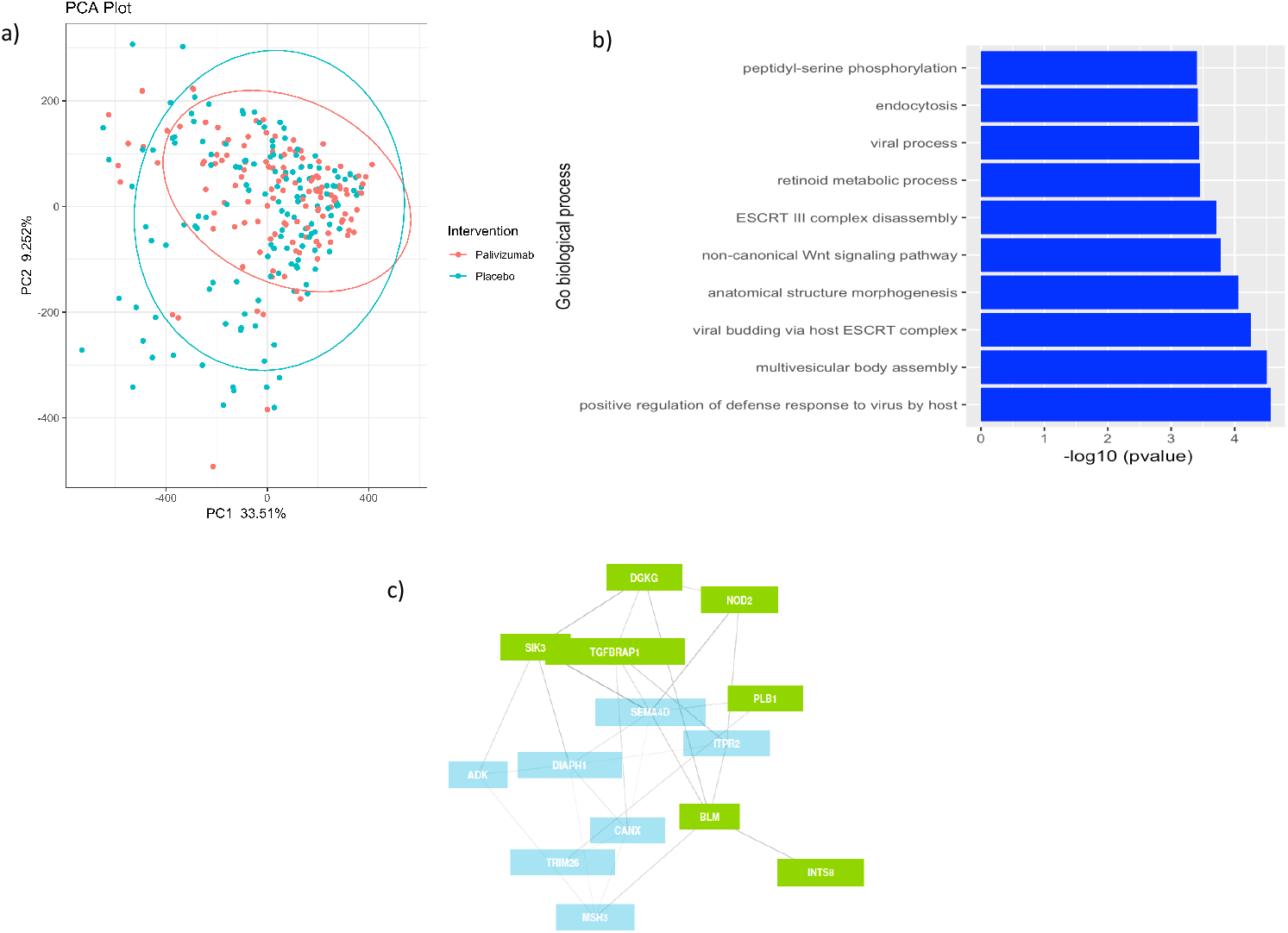
Principal component analysis (PCA) of global methylation patterns. **A**, PCA plot of global methylation patterns in DNA obtained nasal epithelial brushing in relation to Palivizumab intervention. Two PCs were significantly associated with the intervention: PC1 (p=8.1×10^−3^) and PC2 (p=1.0×10^−2^). **B**, Gene set analysis of PC1 using Go-biological process databases. **C**, Gene network of genes that significantly contribute to PC1. These genes annotate to immune related genes that were differentially methylated after RSV immunoprophylaxis in the first year of life. There are two clusters (blue and green) in this network.

**Figure 3:**
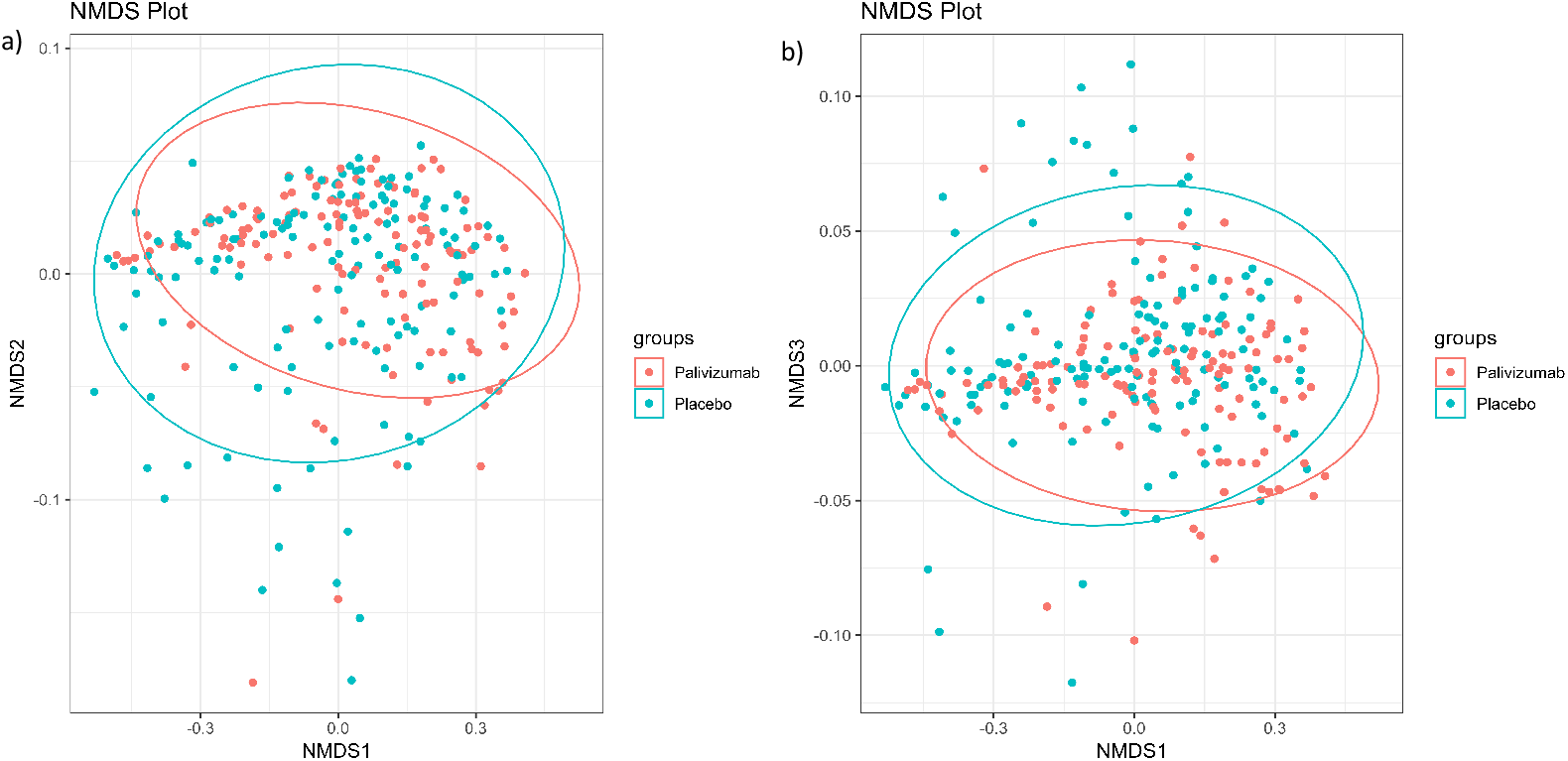
Non metric dimensional biplot. NMDS biplots depicting the individual overall methylation levels (data points, n=274), colored by treatment group: placebo (blue, n=132) and Palivizumab (red, n=142). The difference in overall methylation levels is significant (adonis, R^2^= 0.0195, P value= 0.0115, with 1999 permutation a) NMDS1-NMDS2 b) NMDS1-NMDS3.

We next performed an Epigenome Wide Association Analysis (EWAS), and three CpG sites (cg18040241, cg08243963 and cg19555973) which are localized in Galactosidase *Beta 1 Like 2* (*GLB1L2)* and *SC5DL* (encoding lathosterol oxidase involved in cholesterol biosynthesis) were genome wide significantly associated with RSV immunoprophylaxis versus placebo (Figure 4a, Table E2, *P*<0.05 after FDR correction). These genes have not been previously implicated in viral immune responses or asthma development. The lambda value was 1.123, which indicated no strong inflation in the model after SVA correction (Figure 4b). In differential methylation region (DMR) analysis, we identified 43 DMRs in relation to RSV immunoprophylaxis (Table E3).

**Figure 4.**
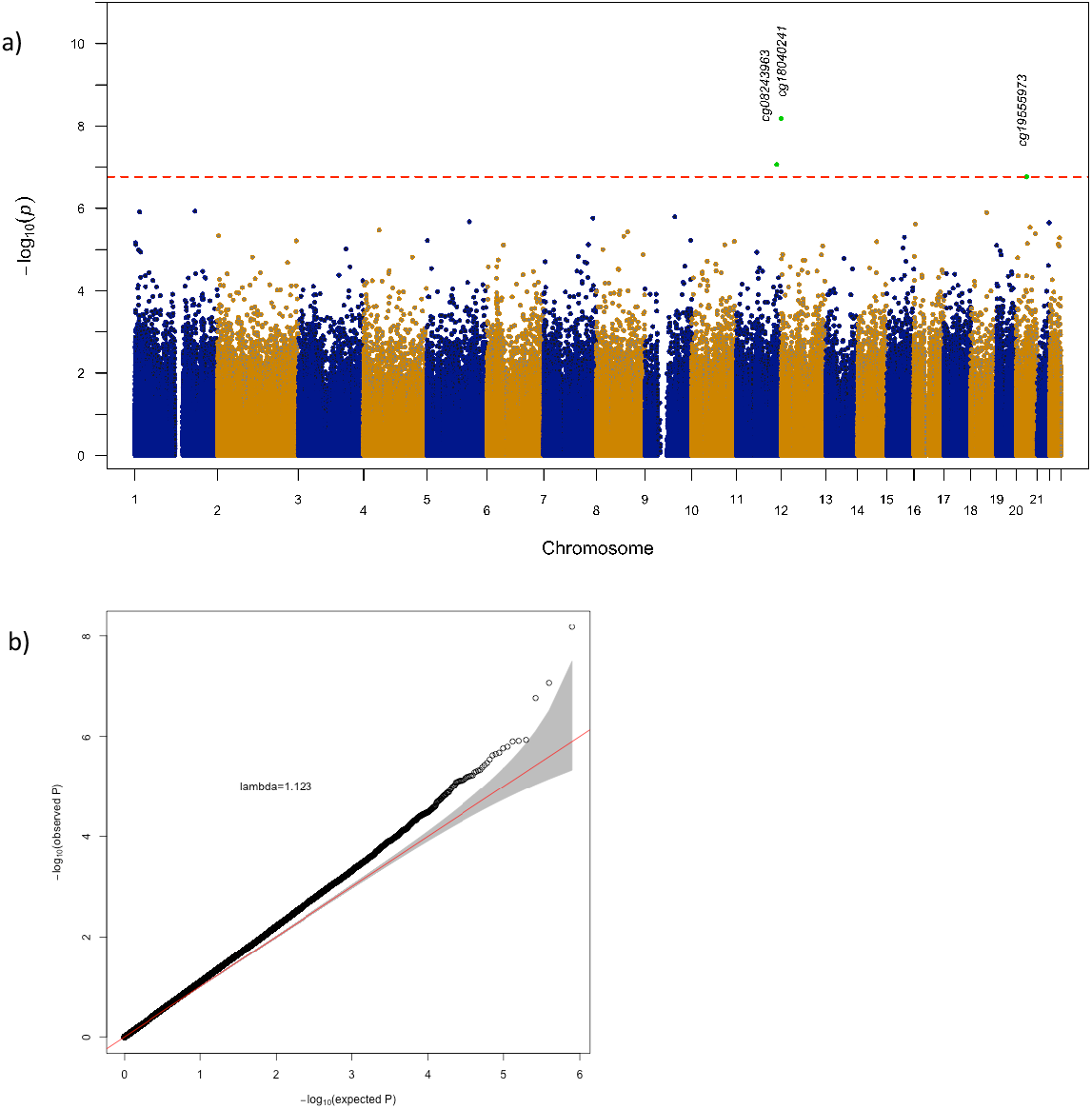
a) Manhattan plot of 790,437 CpG sites for differentially methylation with early life RSV prophylaxis by palivizumab. P values (y axis) correspond to the significance of the difference in methylation between prevention and no prevention subject. The red line correspond to the P value FDR=0.05 threshed. b) A quantile-quantile (QQ) plot displays the experimentally observed Pvalues (vertical axis) as well as the expected Pvalues of a null distribution (horizontal axis). The observed pvalue is obtained from the association between CpG sites methylation and RSV prophylaxis. The model to test the association is methylation ∼ RSV prophylaxis + age at nasal brush sample + sex + maternal smoking + batch, without correction of surrogate variables. The grey area represents the 95% concentration band.

### Pathway analysis

To identify pathways explaining these overall methylation differences after RSV immunoprophylaxis, we selected the top 20 CpG sites contributing most to the first two PCs (Table E4 and E5). We next performed pathway analysis by Gene Network[22] on genes located closely to these CpG sites. The pathway analysis of PC1 indicated enrichment of Go biological pathways involved in “positive regulation of defense response to virus by host” driven by *NOD2* (a known innate immunity gene), *DGKG, MSH3* and *ITPR2*, and “viral budding via host endosomal sorting complexes required for transport (ESCRT)” driven by *CANX, SEMA4D, ITPR2* and *ADK* (Figure 2b, Table E6). Based on gene network clustering, Figure 2c depicts two clusters in this gene network: “positive regulation of interleukin-1 beta secretion” and “antigen processing and presentation of peptide antigen via MHC class I” (Table E7 and E8). The pathway analysis on genes linked to PC2 showed relation to innate immune response genes (Table E9).

### Link of RSV prophylaxis epigenetic markers to asthma and allergy

We next related PC1 of RSV immunoprophylaxis associated methylation to patient reported asthma at age 6 years, but did not observe an association (Point-Biserial correlation −0.077, *P*=0.20). The three CpG sites which were associated with RSV immunoprophylaxis were also not associated with patient reported asthma (Table E10). We linked RSV prevention related epigenetic markers to asthma and allergy associated CpG sites at regional level as many well-known asthma and allergy genes have been identified (Table E3). This differential methylation region analysis identified regions near genes that were previously implicated in the development of asthma and allergy such as *HLA-DPA1, HLA-DPB1, FASLG*, and *CHI3L1*. [24] We further tested 30 CpG sites which were reported to be associated with allergic asthma from a recent nasal epigenomics study[23], yet none of these were associated with RSV immunoprophylaxis (Table E11). However, we did replicate 16 of these 30 CpG sites associated with allergic asthma in nasal epithelium in our study (Table E12).

## Discussion

This work shows that RSV prophylaxis by palivizumab at infancy has global and persistent effects on nasal DNA methylation patterns in childhood. We could observe such changes at global, single CpG site and regional level. Using pathway analysis, we linked these immunoprophylaxis associated global DNA methylation changes to viral response and viral budding genes. We propose that prevention of RSV respiratory tract infection in early childhood leads to different timing and severity of early life RSV respiratory tract infection, which in turn affects local DNA methylation of relevant innate immune response genes. This was further evidenced by differential methylation of cg16834953 in the *NOD2* gene, which functions as a viral pattern recognition receptor and participates in inducing antiviral signaling [24].

An important question is whether the epigenetic patterns we identified would lead to functional differences of these nasal cells, for example in response to other respiratory infections. An alternative explanation would be a direct effect of palivizumab on global gene methylation patterns, or microbiome changes in relation to early life RSV infections that in turn could affect nasal DNA methylation. To our knowledge, this is the first time that long-term methylation effects could be related to the timing of early life RSV infection. We performed our study within the framework of the randomized controlled MAKI trial, enabling us to infer causality rather than reporting an association, as can be done in observational cohorts. As we expect all children to have had RSV respiratory tract infections at the age of six years, we interpret our findings on changes in palivizumab related DNA methylation as an effect of timing and/or severity of early life RSV infection. However, this hypothesis needs further validation in future studies, focused on the timing and severity of early life RSV infection, preferentially in a longitudinal design. The low prevalence of documented RSV infection in the first year of life did not enable us to perform these analyses in this cohort (see table 1).

We did not identify a direct link between the epigenetic markers associated with RSV immunoprophylaxis and asthma at age 6 years in global and single site analyses, but identified some overlap in regional analysis. Although some asthma genes were identified, there was no strong overlap between the RSV immunoprophylaxis associated gene set and previously published asthma associated CpG sites. This is in line with the clinical findings in this cohort, reporting no strong association with early life palivizumab prevention and doctor’s diagnosed asthma at age 6 years[3]. Much larger studies are needed to identify a potential link between RSV immunoprophylaxis, epigenetic changes and asthma[25].

Despite the overall robustness and biological plausibility of our study findings, there are some limitations to consider. Firstly, to the best of our knowledge, there are no suitable data or cohorts available for replication of our findings. The study by O’Brien *et al*.[26] did not include epigenetic studies in the long-term follow up [personal communication]. Second, there is no gold standard to define cell composition in nasal brushes, so we used surrogates variables[18] to represent cell types. Third, our study was single blinded. Fourth, we could not directly confirm that these methylation changes in nasal epithelium regulated gene expression, which should be part of future studies. Fifth, although we studied over 800,00 CpG sites of the human methylome, this still represents about 3 % of all human CpG sites. Further work confirming these differentially methylated sites and detailed assessment of the associated regions using pyrosequencing should be performed. Sixth, other factors may also associate with DNA methylations, such as age[27], cigarette smoke[28], air pollution[29], or other respiratory infections. However, given that our analysis was embedded in a randomized controlled clinical trial, it is very reasonable to assume that potential confounders are distributed equally across the two treatment arms; and that the results were driven by the RSV Immunoprohylaxis.

In conclusion, infant RSV prophylaxis persistently changes nasal epithelial DNA methylation at least until age 6 years, and these methylation profiles are linked to regulation of anti-viral host defense responses and viral budding process.

## Data Availability

The epigenetic datasets generated and analyzed are available from the corresponding author on reasonable request.

## Abbreviations

RSV: Respiratory syncytial virus;
EWAS: Epigenome-wide association studies;
PCA: Principal components analysis;
NMDS: Nonmetric multidimensional scaling;
DMR: Differential methylation region;
ESCRT: Endosomal sorting complexes required for transport.

## Acknowledgements

The authors are indebted to all the participating children and their families for their commitment and participation. We thank all members of the MAKI research team, including the staff of the pediatric lung function laboratory, and the laboratory staff. We also thank the staff of the Genomics Analysis Facility for providing technical assistance with the methylation analyses of this study.

## Authors’ contributions

The study was conceived and designed by LJB and GHK. CJX, CQ and RV performed statistical analyses, CJX drafted the manuscript and interpreted the data. NMS, LJB and GHK contributed to the manuscript writing. NMS, LBCK, EEN, CK and LJB obtained informed consent and clinical information. All authors reviewed and provided comments to the final manuscript (CJX, NMS, CQ, RV, LBCK, EEN, CK, LJB, GHK). All authors read and approved the final manuscript.

## Ethics approval and consent to participate

The MAKI trial was approved by the ethical committee Medisch Ethische Toetsingscommissie (METC) of the Universitair Medisch Centrum Utrecht(21/07/2008), and all study participants provided written consent.

---

**Table E1.** The association of the first ten principal components with Infant RSV immunoprophylaxis.

| Principal components | Associated p value* |
| --- | --- |
| **PC1** | **$8.11 \times 10^{-3}$** |
| **PC2** | **$1.04 \times 10^{-2}$** |
| PC3 | $2.94 \times 10^{-1}$ |
| PC4 | $9.99 \times 10^{-1}$ |
| PC5 | $8.44 \times 10^{-1}$ |
| PC6 | $7.08 \times 10^{-2}$ |
| PC7 | $7.25 \times 10^{-1}$ |
| PC8 | $7.42 \times 10^{-1}$ |
| PC9 | $2.00 \times 10^{-1}$ |
| PC10 | $9.82 \times 10^{-1}$ |

Legend to Table E1: \* Association was investigated with logistic regression model to test the association of RSV immunoprophylaxis ~ principal components).

**Table E2.**
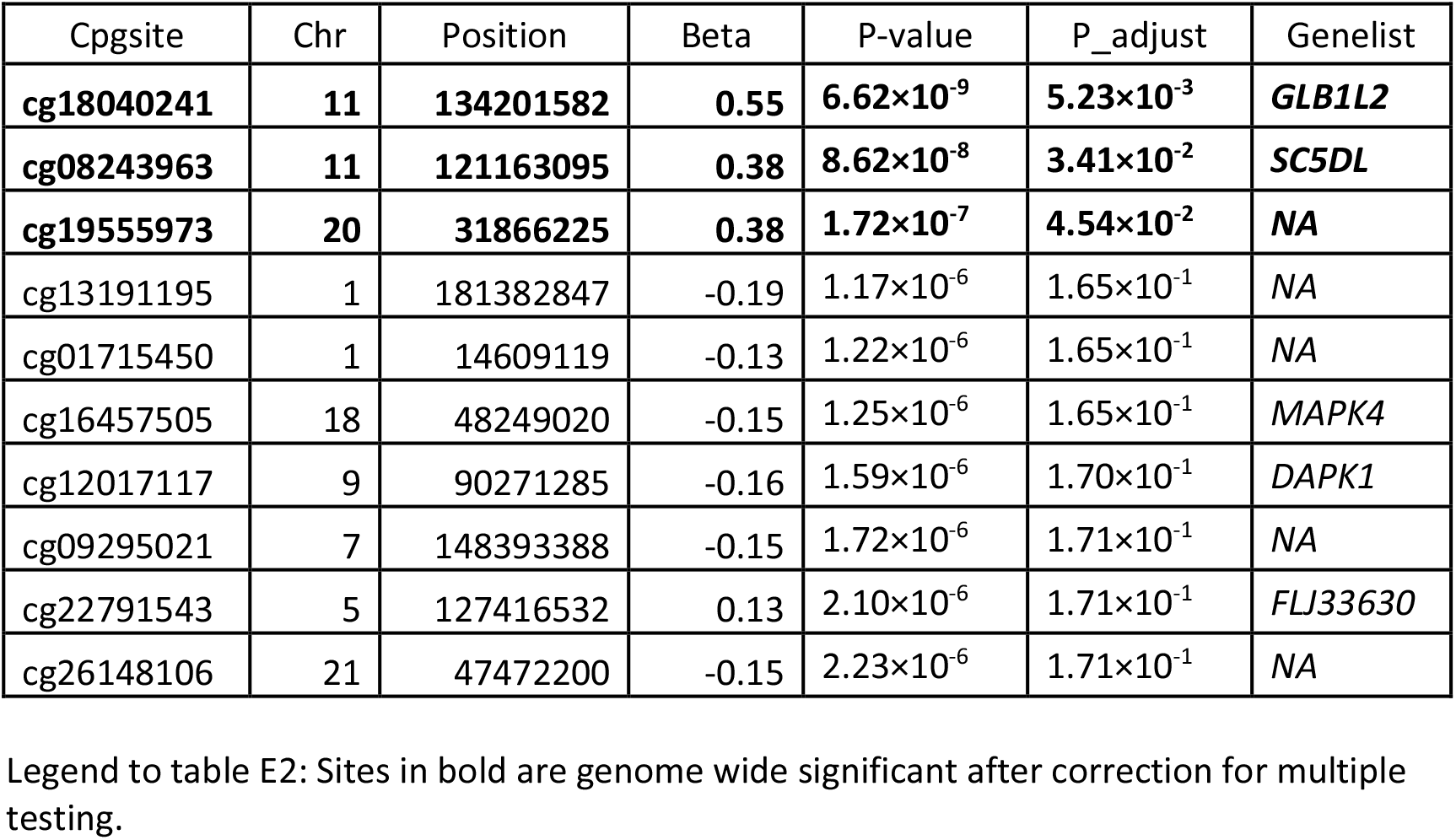
Associations between DNA methylation and RSV immunoprophylaxis with adjustment for surrogate variables.

| Cpgsite | Chr | Position | Beta | P-value | P_adjust | Genelist |
| --- | --- | --- | --- | --- | --- | --- |
| **cg18040241** | **11** | **134201582** | **0.55** | **$6.62 \times 10^{-9}$** | **$5.23 \times 10^{-3}$** | ***GLB1L2*** |
| **cg08243963** | **11** | **121163095** | **0.38** | **$8.62 \times 10^{-8}$** | **$3.41 \times 10^{-2}$** | ***SC5DL*** |
| **cg19555973** | **20** | **31866225** | **0.38** | **$1.72 \times 10^{-7}$** | **$4.54 \times 10^{-2}$** | ***NA*** |
| cg13191195 | 1 | 181382847 | -0.19 | $1.17 \times 10^{-6}$ | $1.65 \times 10^{-1}$ | *NA* |
| cg01715450 | 1 | 14609119 | -0.13 | $1.22 \times 10^{-6}$ | $1.65 \times 10^{-1}$ | *NA* |
| cg16457505 | 18 | 48249020 | -0.15 | $1.25 \times 10^{-6}$ | $1.65 \times 10^{-1}$ | *MAPK4* |
| cg12017117 | 9 | 90271285 | -0.16 | $1.59 \times 10^{-6}$ | $1.70 \times 10^{-1}$ | *DAPK1* |
| cg09295021 | 7 | 148393388 | -0.15 | $1.72 \times 10^{-6}$ | $1.71 \times 10^{-1}$ | *NA* |
| cg22791543 | 5 | 127416532 | 0.13 | $2.10 \times 10^{-6}$ | $1.71 \times 10^{-1}$ | *FLJ33630* |
| cg26148106 | 21 | 47472200 | -0.15 | $2.23 \times 10^{-6}$ | $1.71 \times 10^{-1}$ | *NA* |

Legend to table E2: Sites in bold are genome wide significant after correction for multiple testing.

**Table E3.** DMRs associated with RSV immunoprophylaxis at an SLK-Sidak P value of less than 0.05.

| Chr | Start | End | Num | slk_sidak_p | Gene | Location |
| --- | --- | --- | --- | --- | --- | --- |
| 15 | 81426347 | 81426821 | 9 | $2.30 \times 10^{-10}$ | *CFAP161* | TSS+intron+utr5+cds |
| 6 | 29894050 | 29894254 | 8 | $1.19 \times 10^{-7}$ | *HCG4B* | nc_exon |
| 6 | 33048254 | 33048920 | 19 | $1.31 \times 10^{-7}$ | *HLA-DPA1;HLA-DPB1* | TSS+intron+exon+utr5;intron+cds |
| 11 | 117069780 | 117070047 | 8 | $8.87 \times 10^{-6}$ | *TAGLN;LOC100652768* | TSS+exon+utr5;nc_intron |
| 11 | 94278324 | 94278604 | 7 | $2.59 \times 10^{-5}$ | *LOC105369438;FUT4* | nc_exon;cds |
| 1 | 17614144 | 17614281 | 2 | $3.82 \times 10^{-5}$ | *PADI3* | intergenic |
| 1 | 203155884 | 203156626 | 6 | $4.59 \times 10^{-5}$ | *CHI3L1* | intergenic |
| 11 | 128812804 | 128813009 | 8 | $4.66 \times 10^{-5}$ | *TP53AIP1* | exon+utr5 |
| 19 | 55660514 | 55660871 | 7 | $5.52 \times 10^{-5}$ | *TNNT1* | TSS+intron+exon+utr5 |
| 17 | 79881468 | 79881544 | 5 | $8.07 \times 10^{-5}$ | *MAFG* | intron+utr5 |
| 17 | 79297435 | 79297707 | 4 | $2.23 \times 10^{-4}$ | *LINC00482* | intergenic |
| 2 | 7171869 | 7172098 | 6 | $2.75 \times 10^{-4}$ | *RNF144A* | intron |
| 6 | 29911265 | 29911559 | 5 | $5.82 \times 10^{-4}$ | *HLA-A* | intron+cds |
| 2 | 121498194 | 121498813 | 3 | $6.26 \times 10^{-4}$ | *GLI2* | intergenic |
| 12 | 89748726 | 89749081 | 8 | $7.97 \times 10^{-4}$ | *DUSP6* | intergenic |
| 8 | 74282708 | 74282932 | 4 | $1.44 \times 10^{-3}$ | *RDH10-AS1* | intergenic |
| 12 | 122019006 | 122019118 | 6 | $1.92 \times 10^{-3}$ | *KDM2B* | intergenic |
| 12 | 130976379 | 130976552 | 3 | $1.94 \times 10^{-3}$ | *RIMBP2* | intron+utr5 |
| 5 | 172751061 | 172751409 | 5 | $2.32 \times 10^{-3}$ | *STC2* | intron |
| 10 | 135342413 | 135342937 | 3 | $2.50 \times 10^{-3}$ | *CYP2E1* | intron |
| 7 | 105319558 | 105319820 | 5 | $4.04 \times 10^{-3}$ | *ATXN7L1* | TSS+exon+utr5;intron |
| 1 | 172608566 | 172608705 | 3 | $4.32 \times 10^{-3}$ | *FASLG* | intergenic |
| 1 | 59042931 | 59043256 | 7 | $4.36 \times 10^{-3}$ | *TACSTD2* | TSS+exon |
| 17 | 38498816 | 38499097 | 4 | $5.99 \times 10^{-3}$ | *RARA;RARA-AS1;RARA* | intron;nc_intron+nc_exon;utr5+cds |
| 4 | 681086 | 681155 | 3 | $6.77 \times 10^{-3}$ | *SLC49A3* | intron |
| 2 | 6072744 | 6072982 | 6 | $7.83 \times 10^{-3}$ | *SILC1* | nc_exon |
| 9 | 96715047 | 96715257 | 2 | $8.39 \times 10^{-3}$ | *BARX1* | intron+cds |
| 6 | 30079139 | 30079281 | 8 | $9.66 \times 10^{-3}$ | *TRIM31-AS1;TRIM31;TRIM31* | nc_intron;nc_intron;intron |
| 8 | 120685124 | 120685318 | 4 | $1.01 \times 10^{-2}$ | *ENPP2* | intron |
| 16 | 70780647 | 70780948 | 4 | $1.24 \times 10^{-2}$ | *VAC14* | intron |
| 7 | 157368881 | 157369159 | 6 | $1.55 \times 10^{-2}$ | *PTPRN2* | intron |
| 19 | 15530606 | 15530871 | 4 | $2.28 \times 10^{-2}$ | *AKAP8L* | intergenic |
| 7 | 4228775 | 4228949 | 2 | $2.42 \times 10^{-2}$ | *SDK1* | intron |
| 22 | 29470209 | 29470348 | 4 | $3.35 \times 10^{-2}$ | *KREMEN1* | intron |
| 6 | 31166502 | 31166800 | 4 | $3.48 \times 10^{-2}$ | *HCG27* | nc_intron |
| 21 | 43735722 | 43735936 | 6 | $3.50 \times 10^{-2}$ | *TFF3* | intergenic |
| 19 | 1155030 | 1155226 | 4 | $3.53 \times 10^{-2}$ | *SBNO2* | intron+utr5 |
| 10 | 31146954 | 31147089 | 2 | $3.82 \times 10^{-2}$ | *ZNF438* | intron;intron+utr5;<br>nc_intron |
| 3 | 52877048 | 52877272 | 3 | $4.04 \times 10^{-2}$ | *STIMATE;STIMATE-MUSTN1* | intron;intron |
| 6 | 15401024 | 15401068 | 3 | $4.17 \times 10^{-2}$ | *JARID2* | intron;intron+utr5 |
| 1 | 108235664 | 108235771 | 2 | $4.24 \times 10^{-2}$ | *VAV3* | intron |
| 9 | 117692745 | 117693072 | 7 | $4.56 \times 10^{-2}$ | *TNFSF8* | TSS+exon+utr5 |
| 22 | 36500566 | 36500794 | 3 | $4.81 \times 10^{-2}$ | *APOL3* | intergenic |

Legend to Table E3:
Chr: Chromosome
Start: start position of the region
End: end position of the region
Num: number of probes in the region
Sidak p-value: adjusted p value by comb-p method

**Table E4.**
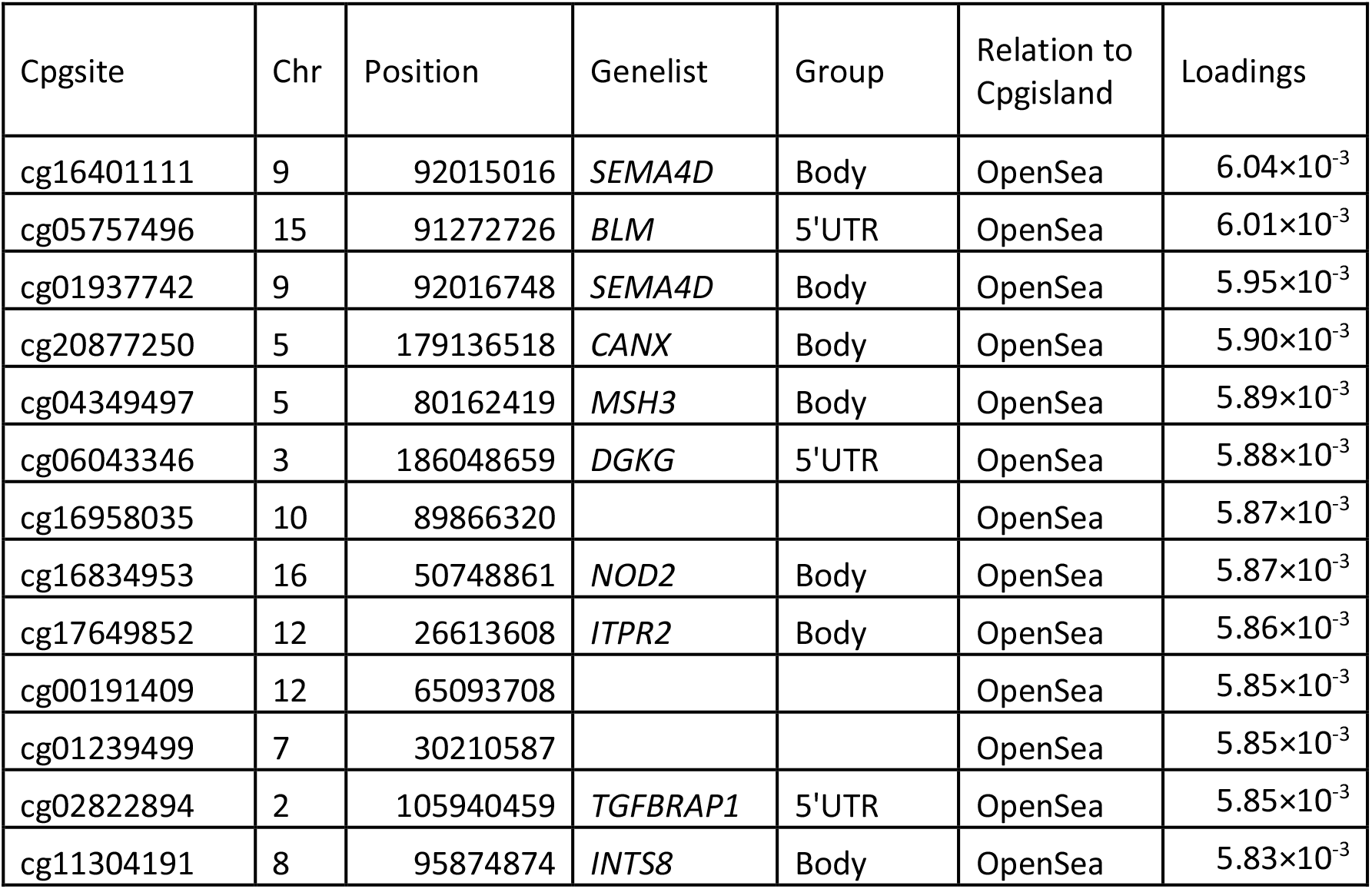
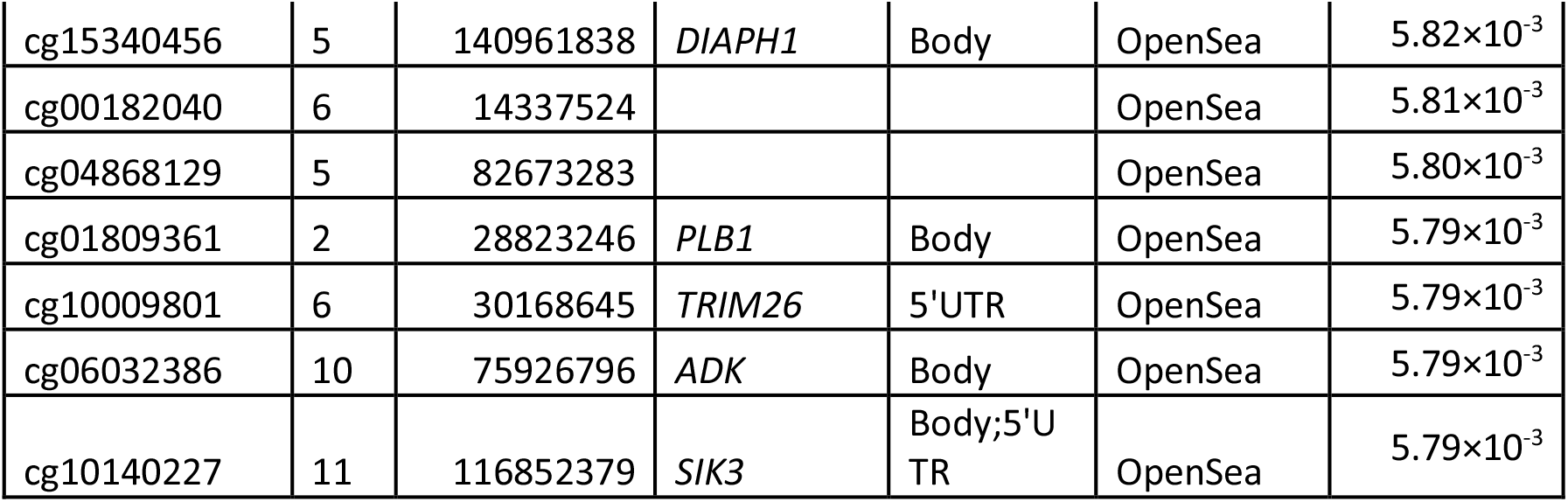
The top 20 CpG sites that contributed most in loadings to the first principal component (PC1).

**Table E5.** The top 20 CpGs that contributed most in loadings to the first principal component (PC2).

| Cpgsite | Chr | Position | Genelist | Group | Relation to Cpg Island | Loadings |
| --- | --- | --- | --- | --- | --- | --- |
| cg01697719 | 22 | 19754125 | *TBX1* | Body | Island | $7.15 \times 10^{-3}$ |
| cg08382235 | 22 | 19754251 | *TBX1* | Body | Island | $6.78 \times 10^{-3}$ |
| cg13332774 | 10 | 106028628 | *GSTO2* | TSS200 | Island | $6.62 \times 10^{-3}$ |
| cg22413388 | 22 | 46367617 | *WNT7B* | Body | Island | $6.62 \times 10^{-3}$ |
| cg17121176 | 6 | 107956004 | *SOBP* | Body | Island | $6.23 \times 10^{-3}$ |
| cg10332422 | 13 | 114292552 | *TFDP1* | Body | Island | $5.62 \times 10^{-3}$ |
| cg10927841 | 2 | 26407743 | *FAM59B* | Body | Island | $5.55 \times 10^{-3}$ |
| cg24477102 | 8 | 2879917 | *CSMD1* | Body | Island | $5.53 \times 10^{-3}$ |
| cg25389087 | 18 | 74824413 | *MBP* | 5'UTR | Island | $5.51 \times 10^{-3}$ |
| cg18456523 | 5 | 54516805 | | | Island | $5.51 \times 10^{-3}$ |
| cg10448954 | 16 | 89593394 | *SPG7* | Body | Island | $5.36 \times 10^{-3}$ |
| cg15894683 | 20 | 56805751 | | | S_Shore | $5.34 \times 10^{-3}$ |
| cg19563510 | 17 | 79881483 | *MAFG* | 5'UTR;TSS200 | S_Shore | $5.34 \times 10^{-3}$ |
| cg11875917 | 8 | 4980497 | | | Island | $5.31 \times 10^{-3}$ |
| cg26900073 | 12 | 132276572 | *SFSWAP* | Body | Island | $5.31 \times 10^{-3}$ |
| cg15271348 | 13 | 114292740 | *TFDP1* | Body | Island | $5.29 \times 10^{-3}$ |
| cg02833580 | 12 | 65996465 | *LOC100507065* | Body | OpenSea | $-5.35 \times 10^{-3}$ |
| cg03577157 | 5 | 80716263 | *RNU510;S*<br>*SBP2* | Body;3'UTR | OpenSea | $-5.45 \times 10^{-3}$ |
| cg23282852 | 6 | 52255402 | *PAQR8* | 5'UTR | OpenSea | $-5.47 \times 10^{-3}$ |
| ch.22.909671F | 22 | 46114168 | *ATXN10* | Body | OpenSea | $-5.76 \times 10^{-3}$ |

**Table E6:**
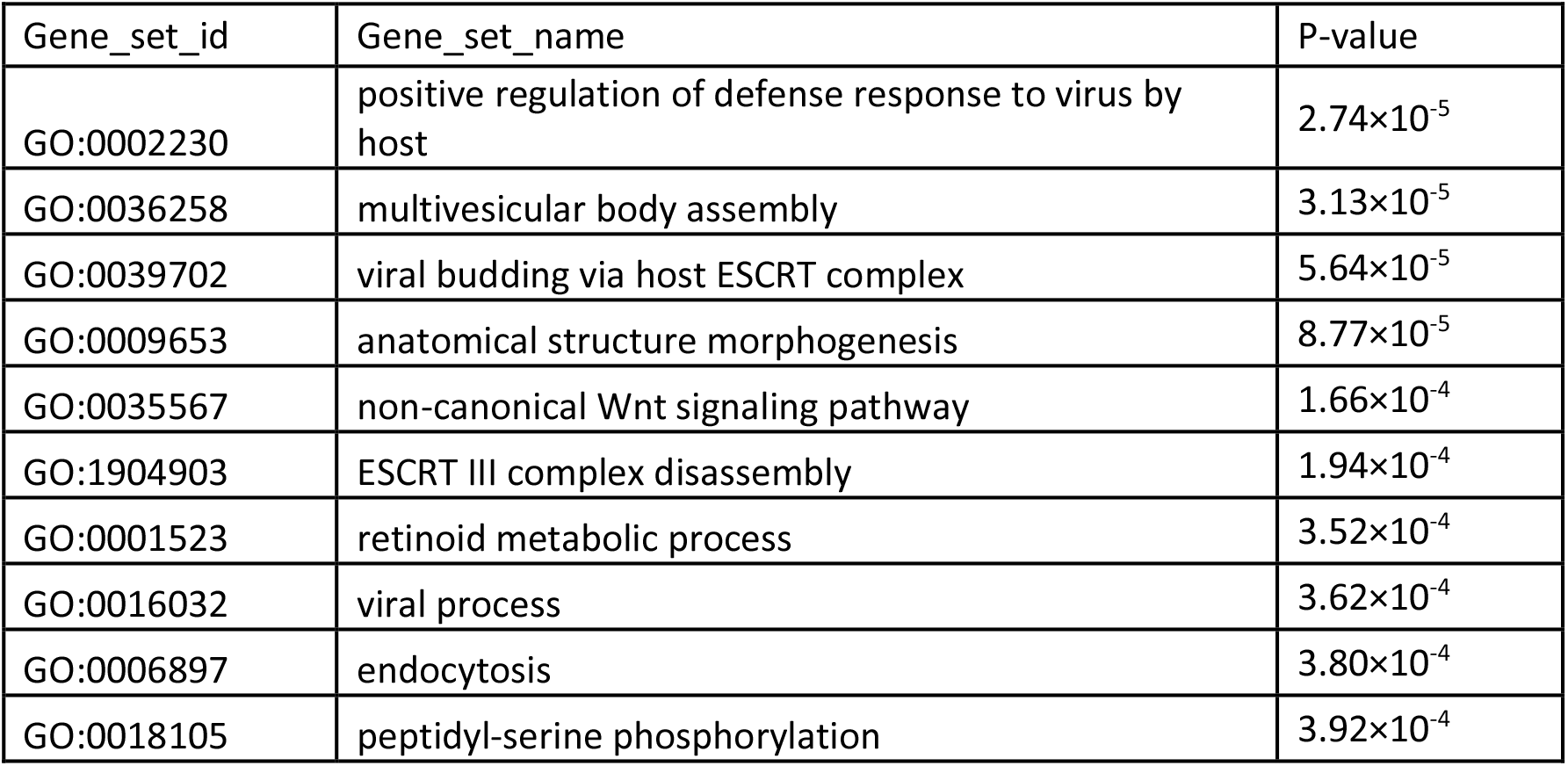
Gene set analysis of PC1 using Go-Biological process databases.

**Table E7:**
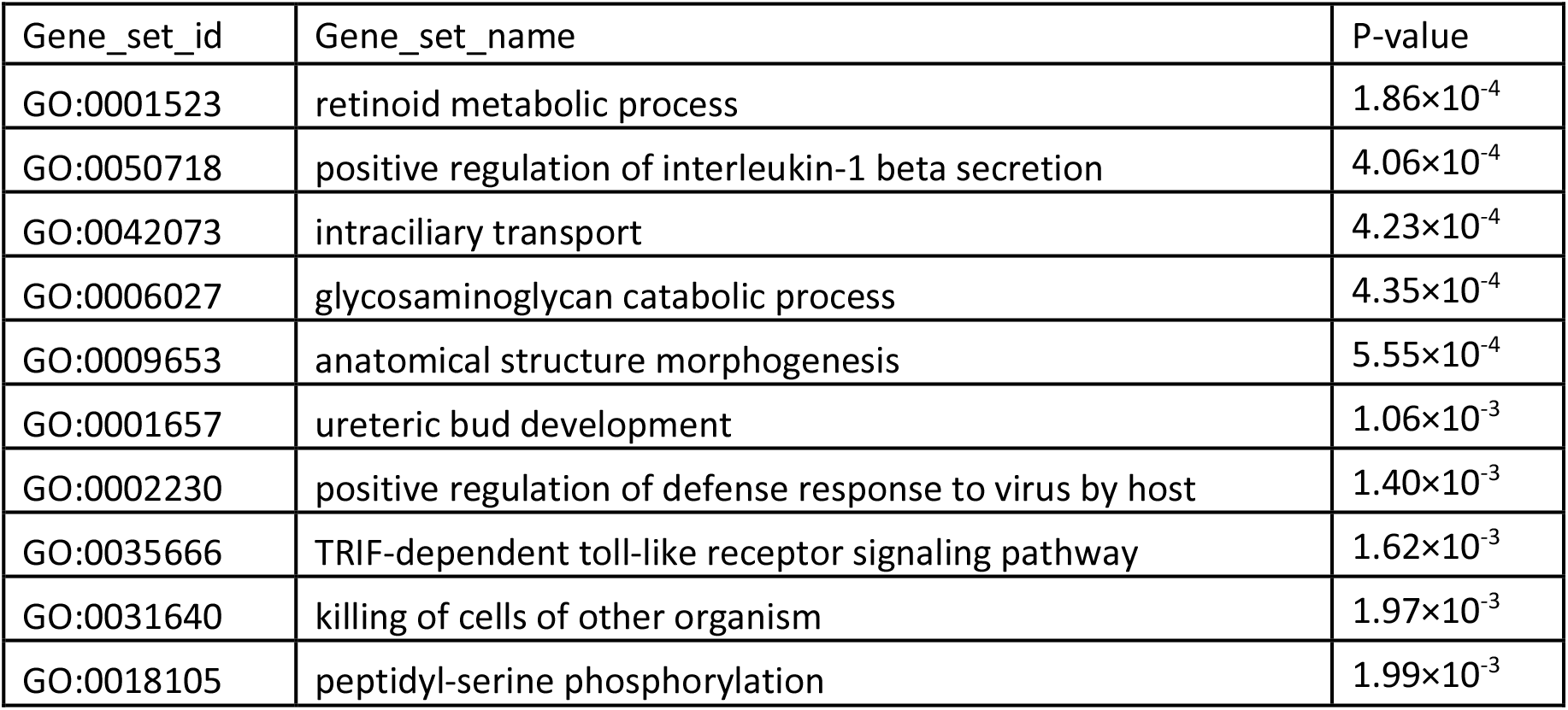
Gene set analysis by using Go-Biological process databases of green cluster of Figure 2c.

**Table E8:** Gene set analysis by using Go-Biological process databases of blue cluster of Figure 2c.

| Gene_set_id | Gene_set_name | P-value |
| --- | --- | --- |
| GO:0006491 | N-glycan processing | $2.09 \times 10^{-5}$ |
| GO:0002474 | antigen processing and presentation of peptide antigen via MHC class I | $4.51 \times 10^{-5}$ |
| GO:0036258 | multivesicular body assembly | $2.19 \times 10^{-4}$ |
| GO:0034975 | protein folding in endoplasmic reticulum | $2.82 \times 10^{-4}$ |
| GO:0048008 | platelet-derived growth factor receptor signaling pathway | $2.90 \times 10^{-4}$ |
| GO:0007064 | mitotic sister chromatid cohesion | $3.79 \times 10^{-4}$ |
| GO:0006665 | sphingolipid metabolic process | $3.79 \times 10^{-4}$ |
| GO:0006626 | protein targeting to mitochondrion | $4.22 \times 10^{-4}$ |
| GO:0039702 | viral budding via host ESCRT complex | $5.46 \times 10^{-4}$ |
| GO:0006699 | bile acid biosynthetic process | $6.33 \times 10^{-4}$ |

**Table E9:** Gene set analysis of PC2 by using Go-Biological process databases.

| Gene_set_id | Gene_set_name | P-value |
| --- | --- | --- |
| GO:0061077 | chaperone-mediated protein folding | $3.92 \times 10^{-4}$ |
| GO:0099132 | ATP hydrolysis coupled cation transmembrane transport | $5.77 \times 10^{-4}$ |
| GO:0045087 | innate immune response | $1.61 \times 10^{-4}$ |
| GO:0002224 | toll-like receptor signaling pathway | $1.79 \times 10^{-3}$ |
| GO:0030317 | flagellated sperm motility | $1.87 \times 10^{-3}$ |
| GO:0042340 | keratan sulfate catabolic process | $2.29 \times 10^{-3}$ |
| GO:0045893 | positive regulation of transcription, DNA-templated | $2.45 \times 10^{-3}$ |
| GO:0071577 | zinc II ion transmembrane transport | $3.55 \times 10^{-3}$ |
| GO:0042752 | regulation of circadian rhythm | $3.74 \times 10^{-3}$ |
| GO:0050829 | defense response to Gram-negative bacterium | $4.15 \times 10^{-3}$ |

**Table E10:** Associations between 3 CpG sites associated with RSV immunoprophylaxis with patient reported asthma.

| Cpgsite | Chr | Position | Effect | P-value | Genelist |
| --- | --- | --- | --- | --- | --- |
| cg18040241 | 11 | 134201582 | -0.40 | 0.08 | *GLB1L2* |
| cg08243963 | 11 | 121163095 | -0.024 | 0.94 | *SC5DL* |
| cg19555973 | 20 | 31866225 | -0.097 | 0.73 | NA |

**Table E11:** Associations between 30 CpG sites, which are reported to associate with allergic asthma in a previous study (Forno et al) with RSV immunoprophylaxis.

| CpG | Gene | Chr | Position | Effect-discovery | Coef | P-value |
| --- | --- | --- | --- | --- | --- | --- |
| cg07239613 | *CES4A* | 16 | 67051005 | -4.83 | 0.05 | 0.07 |
| cg00664723 | *FBXL7* | 5 | 15927184 | -1.81 | -0.06 | 0.08 |
| cg01870976 | *PCSK6* | 15 | 101887154 | -1.79 | 0.05 | 0.19 |
| cg04132353 | *CAPN14* | 2 | 31440349 | -1.91 | 0.04 | 0.27 |
| cg20790648 | *SUCNR1* | 3 | 151619923 | -1.87 | 0.05 | 0.27 |
| cg24707200 | *NTRK1* | 1 | 156833163 | -4.91 | -0.02 | 0.32 |
| cg22855021 | *TSHR* | 14 | 81610812 | -1.63 | 0.05 | 0.33 |
| cg10549071 | *SPP2* | 2 | 235160451 | -2.3 | 0.03 | 0.37 |
| cg10099207 | *C15orf54* | 15 | 39544144 | -1.4 | -0.05 | 0.40 |
| cg01859321 | *PLEC* | 8 | 144970195 | -1.45 | 0.03 | 0.45 |
| cg15006973 | *GJA4* | 1 | 35258933 | -2.86 | -0.02 | 0.48 |
| cg06675531 | *SYNPO* | 5 | 150019123 | -4.4 | -0.02 | 0.52 |
| cg09472600 | *NCF2* | 1 | 183537770 | -1.65 | 0.03 | 0.55 |
| cg21158502 | *ANKRD31* | 5 | 74348187 | -2.16 | -0.02 | 0.55 |
| cg16518142 | *CDH26* | 20 | 58533713 | -2.89 | -0.01 | 0.61 |
| cg18749617 | *PCSK6* | 15 | 102028637 | -1.49 | -0.01 | 0.68 |
| cg13586696 | *C22orf31* | 22 | 29458723 | -1.63 | -0.02 | 0.71 |
| cg13570892 | *DUOX1* | 15 | 45449437 | -1.61 | 0.02 | 0.71 |
| cg22862094 | *EFNA5* | 5 | 106859865 | -2.07 | -0.01 | 0.71 |
| cg16027132 | *CDHR3* | 7 | 105521116 | -1.6 | 0.01 | 0.76 |
| cg19497511 | *LRRFIP1* | 2 | 238609807 | -3.44 | -0.01 | 0.76 |
| cg08175352 | *ZPLD1* | 3 | 101894206 | -1.72 | -0.02 | 0.78 |
| cg00406211 | *GRK5* | 10 | 121077022 | -2 | 0.01 | 0.78 |
| cg17223698 | *C15orf54* | 15 | 39416631 | -3.31 | 0.00 | 0.88 |
| cg19610615 | *ADCK1* | 14 | 78446340 | -1.6 | 0.00 | 0.90 |
| cg20372759 | *METTL1* | 12 | 58162287 | -1.97 | 0.00 | 0.91 |
| cg03387497 | *BANF2* | 20 | 17680945 | -1.88 | 0.00 | 0.97 |
| cg03875819 | *LINC00704* | 10 | 4386802 | -1.65 | 0.00 | 0.98 |
| cg10830021 | *NUP98* | 11 | 3815589 | -1.5 | 0.00 | 0.99 |
| cg08844313 | *PDE6A* | 5 | 149240529 | -1.92 | NA | NA |

**Table E12:** Associations between 30 CpG sites, which are reported to associate with allergic asthma with allergic asthma in MAKI data and 16 out of 29 could be replicated (P value<0.05).

| CpG | Gene | Chr | Position | Effect-discovery* | Effect-Maki | P-value |
| --- | --- | --- | --- | --- | --- | --- |
| cg20372759 | *METTL1* | 12 | 58162287 | -1.97 | -6.10 | **3.53 ×10<sup>-4</sup>** |
| cg15006973 | *GJA4* | 1 | 35258933 | -2.86 | -5.99 | **3.82 ×10<sup>-4</sup>** |
| cg24707200 | *NTRK1* | 1 | 156833163 | -4.91 | -6.78 | **1.22 ×10<sup>-3</sup>** |
| cg09472600 | *NCF2* | 1 | 183537770 | -2.16 | -2.84 | **1.80 ×10<sup>-3</sup>** |
| cg10830021 | *NUP98* | 11 | 3815589 | -1.5 | -4.94 | **2.27 ×10<sup>-3</sup>** |
| cg22855021 | *TSHR* | 14 | 81610812 | -1.63 | -2.03 | **5.16 ×10<sup>-3</sup>** |
| cg10549071 | *SPP2* | 2 | 235160451 | -2.3 | -2.93 | **6.27 ×10<sup>-3</sup>** |
| cg10099207 | *C15orf54* | 15 | 39544144 | -1.4 | -2.01 | **1.16 ×10<sup>-2</sup>** |
| cg08175352 | *ZPLD1* | 3 | 101894206 | -1.72 | -2.26 | **1.43 ×10<sup>-2</sup>** |
| cg20790648 | *SUCNR1* | 3 | 151619923 | -1.87 | -2.12 | **1.53 ×10<sup>-2</sup>** |
| cg00664723 | *FBXL7* | 5 | 15927184 | -1.81 | -3.48 | **1.56 ×10<sup>-2</sup>** |
| cg16518142 | *CDH26* | 20 | 58533713 | -2.89 | -4.49 | **1.67 ×10<sup>-2</sup>** |
| cg18749617 | *PCSK6* | 15 | 102028637 | -1.49 | -2.30 | **2.31 ×10<sup>-2</sup>** |
| cg01870976 | *PCSK6* | 15 | 101887154 | -1.79 | -2.33 | **3.26 ×10<sup>-2</sup>** |
| cg00406211 | *GRK5* | 10 | 121077022 | -2 | -2.92 | **4.26 ×10<sup>-2</sup>** |
| cg19497511 | *LRRFIP1* | 2 | 238609807 | -3.44 | -3.41 | **4.81 ×10<sup>-2</sup>** |
| cg17223698 | *C15orf54* | 15 | 39416631 | -1.6 | -1.98 | 5.01 ×10<sup>-2</sup> |
| cg19610615 | *ADCK1* | 14 | 78446340 | -3.31 | -2.37 | 5.45 ×10<sup>-2</sup> |
| cg22862094 | *EFNA5* | 5 | 106859865 | -2.07 | -1.58 | 6.33 ×10<sup>-2</sup> |
| cg03875819 | *LINC00704* | 10 | 4386802 | -1.65 | -1.37 | 1.73 ×10<sup>-1</sup> |
| cg03387497 | *BANF2* | 20 | 17680945 | -1.88 | -1.49 | 1.73 ×10<sup>-1</sup> |
| cg21158502 | *ANKRD31* | 5 | 74348187 | -1.65 | -1.02 | 2.28 ×10<sup>-1</sup> |
| cg13586696 | *C22orf31* | 22 | 29458723 | -1.63 | -1.24 | 2.31 ×10<sup>-1</sup> |
| cg01859321 | *PLEC* | 8 | 144970195 | -1.45 | -1.00 | 2.71 ×10<sup>-1</sup> |
| cg16027132 | *CDHR3* | 7 | 105521116 | -1.6 | -0.78 | 4.06 ×10<sup>-1</sup> |
| cg13570892 | *DUOX1* | 15 | 45449437 | -1.61 | -0.53 | 5.02 ×10<sup>-1</sup> |
| cg06675531 | *SYNPO* | 5 | 150019123 | -4.4 | -0.80 | 5.98 ×10<sup>-1</sup> |
| cg04132353 | *CAPN14* | 2 | 31440349 | -1.91 | -0.34 | 7.10 ×10<sup>-1</sup> |
| cg07239613 | *CES4A* | 16 | 67051005 | -4.83 | 0.07 | 9.51 ×10<sup>-1</sup> |
| cg08844313 | *PDE6A* | 5 | 149240529 | -1.92 | NA | NA |

